# Participation Questionnaire for Preschoolers with Autism Spectrum Disorder: Item Development

**DOI:** 10.1101/2023.08.22.23294206

**Authors:** Takuto Nakamura, Sakumi Koyama, Hirofumi Nagayama, Satoshi Sasada

## Abstract

Occupational therapists need to comprehensively assess participation of children with autism spectrum disorder (ASD) in daily activities and evaluate the effectiveness of relevant interventions. Several participation measurement tools have been developed for children with ASD, but these tools require expert involvement, which is a barrier to large-scale surveys. To address these concerns, a caregiver-administered questionnaire—the Participation Questionnaire for Preschoolers (PQP)—was developed. However, this tool could be improved due to its narrow age range of 48–72 months and because the item development process does not reflect the perspectives of children and caregivers. Therefore, we expanded the PQP’s target age range to 36–83 months and developed new items that reflect the perspectives of professionals and caregivers. Interviews were conducted with eight experts in supporting children with ASD and 11 caregivers of children with ASD. The interviews were transcribed, and a content analysis was performed. The number of questions was reduced from 51 to 36, and the order of items was changed for clarity. Two of the eight sub-domains were removed to clarify the conceptual difference between activity and participation. The updated version of the PQP has two unique features: (1) it can be administered without expert involvement, and (2) it includes items specific to the challenges faced by children with ASD. Future development of the scale and validation of its measurement properties are needed.

## Introduction

Autism spectrum disorder (ASD) is a neurodevelopmental disorder characterized by challenges related to social interaction, communication, and repetitive behaviors and interests [1]. Typically, ASD is evident early in an individual’s development, particularly in childhood, and can considerably impact later life. Therefore, support in early childhood is vital, and significant financial resources are invested in high-income countries to manage ASD [2].

“Participation” is a construct defined in the International Classification of Functioning, Disability, and Health as “involvement in life situations” [3]. This construct, which implies involvement in academics, family roles, interactions with friends, and community activities, is an important goal for occupational therapists [4]. However, the disability characteristics of ASD create limitations to this participation [5]. Prior research suggests that children with ASD have limited participation in many domains compared to typically developing children, and that this limitation is more prominent in ASD than in other disorders [6, 7].

Occupational therapists have examined the use of standardized methods to assess the participation of children with ASD [8]. Many of the earliest participation measurement tools were generic tools not limited to a specific disability or disorder. These tools have helped to characterize participation across disability status and type and build a knowledge base for participation-focused occupational therapy practice. However, in recent years, there has been a rapid expansion in the development of disability-specific participation measurement tools that are highly useful in clinical settings and aptly reflect the effects of interventions. For example, tools such as the “Activities Scale for Kids performance (ASKp) [9]” for measuring participation in children with musculoskeletal disorders, and “Pediatric Measure of Participation short forms [10]” for children with spinal cord injuries, have been developed and utilized for various disabilities. This trend emphasizes the need to develop disability-specific participation measurement tools for children with ASD [8].

The oldest existing disability-specific participation measurement tool for children with ASD is the Matrix for Assessment of Activities and Participation [11]. This tool can be used by professionals to comprehensively measure the activities and participation of children with ASD aged 3–6 years. The Structured Preschool Participation Observation for Children with ASD (SPO-ASD) can specifically measure participation in non-inclusive educational settings [12]. The SPO-ASD is a valid and reliable tool designed for professionals to conduct detailed assessments of ASD features in children aged 4-6 years. Additionally, the Picture My Participation-Chinese version is a valid tool that can be used for children with ASD [13]. This tool is advantageous because it allows children to self-report their participation through a photo-based interview and can be used by children with ASD aged 5–9 years.

However, the use of these measurement tools necessitates trained experts to conduct interviews or observations. These requirements pose challenges not only for clinical application [14] but also for conducting large-scale surveys.

Given the constraints of these previously developed tools, the Participation Questionnaire for Preschoolers (PQP) was developed [15]. The PQP, developed through interviews with caregivers and professionals, is a participation measurement tool specifically designed for children with ASD. It is a caregiver-administered questionnaire comprising 54 items that comprehensively measure participation and can be answered without professional involvement. The development of a simple, standardized measurement tool such as the PQP can help clinicians easily screen for and monitor changes in participation. Furthermore, the questionnaire can serve as a basis for building evidence in support of participation of children with ASD in large-scale surveys.

However, development of the PQP items is ongoing. Furthermore, the target age range is limited to 48–72 months. Considering that ASD is often detected at approximately three years of age in high-income countries [2], the PQP’s current target age range may not satisfy the needs of actual clinical practice and research. There is also a growing demand for new tools to assess and validate changes in participation and the effectiveness of interventions for children with ASD. Therefore, this study aims to expand the PQP’s target age range and develop new items adaptable to children of diverse age groups.

## Materials and methods

We used a qualitative methodology to develop the questionnaire items. To ensure that the constructs captured important indicators related to the participation of children with ASD, the item development steps of the COnsensus-based Standards for the selection of health Measurement Instruments (COSMIN) methodology for assessing the content validity of patient-reported outcome measures (PROMs)-user manual were used as a reference [16]. Three main steps were followed to develop the questionnaire. First, we determined the basic design of the questionnaire, including the constructs to be measured, the target population, and the response options. This study followed the PQP design used in previous studies [15], except for the target age group. Next, content experts (described below) reviewed the 54 items of the PQP [15] and participated in interviews to assess content validity. Semi-structured interviews were conducted to collect the perspectives of caregivers of children with ASD. Additionally, cognitive interviews were conducted with caregivers to judge whether the items were interpreted as the developers intended. The institutional review board of the first author’s university approved the study, and all participants provided informed consent. This study was conducted between February and October 2021. It was, however, not prospectively registered in any public open access database.

### Validation by experts

#### Questionnaire design

The PQP is based on the International Classification of Functioning (ICF), a comprehensive framework developed by the World Health Organization (WHO) [3] to assess health and disability [15]. The PQP was developed for professionals to assess the participation of children with ASD in real-life situations and provide targeted interventions. A reflective model is adopted in the PQP, and a diverse range of indicators is established to capture the overall picture of “participation.” The preliminary PQP items were developed in Japanese and included eight tentative sub-areas based on the International Classification of Functioning, Children, and Youth (ICF-CY) codes related to activity and participation. These sub-domains are communication; mobility; self-care; domestic life; interpersonal interactions and relationships; engagement in play; education; and community, social, and civic life [3]. To evaluate participation in these eight domains, the PQP included both the dimensions of performance and capacity, as defined by the ICF. Furthermore, to facilitate a more comprehensive discussion of participation dimensions during interviews, the initial items incorporated elements such as a child’s enjoyment, frequency of engagement, degree of involvement, and parental aspirations, based on findings from literature reviews [17, 18, 19]. This approach was intended to ensure that each item reflects various aspects of participation, ultimately contributing to a comprehensive representation of participation as a whole.

Interviews were conducted with experts with at least five years of experience working with children with ASD to validate the questionnaire content. We gathered feedback from four occupational therapists, three researchers in occupational therapy theory, one “participation” researcher, one physician, one speech and hearing therapist, one clinical psychologist, one certified social worker, one childcare worker, and one physical therapist. All participants had clinical experience in Japan. Based on their input, a 51-item draft of the questionnaire was developed (Table 1). Response options were set on a 5-point scale: “agree” (5 points), “somewhat agree” (4 points), “undecided” (3 points), “somewhat disagree” (2 points), and “disagree” (1 point). The total score was calculated by summing the scores for each item. The target population for the PQP is preschool-aged ASD children with an intelligence quotient (IQ) of 50 or higher who have no comorbidities other than neurodevelopmental disorders [20]. Although the PQP’s target age range was 48–72 months, in this study, the items were modified to verify the content validity when the target age range was expanded to 36–83 months.

**Table 1.**
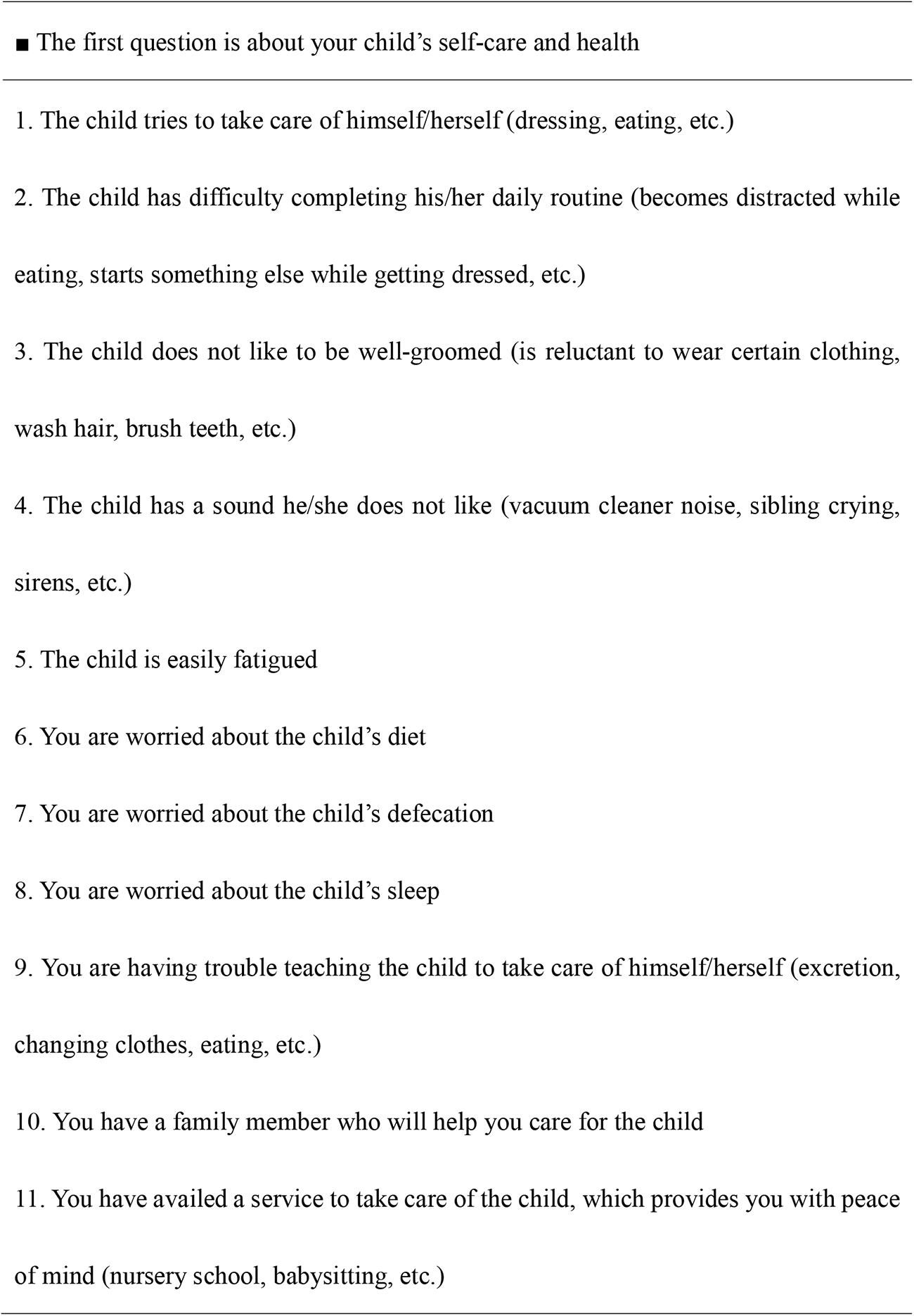
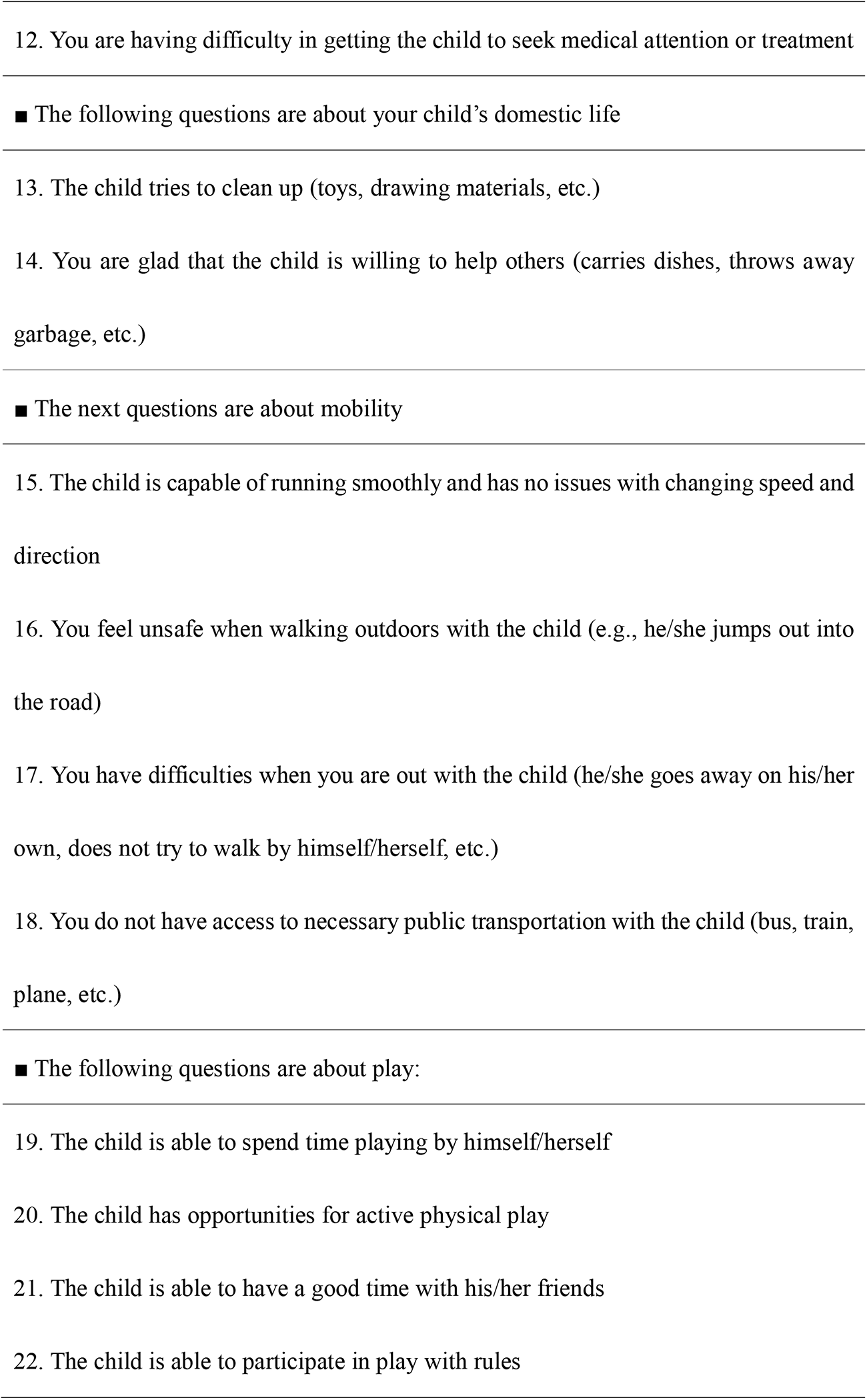
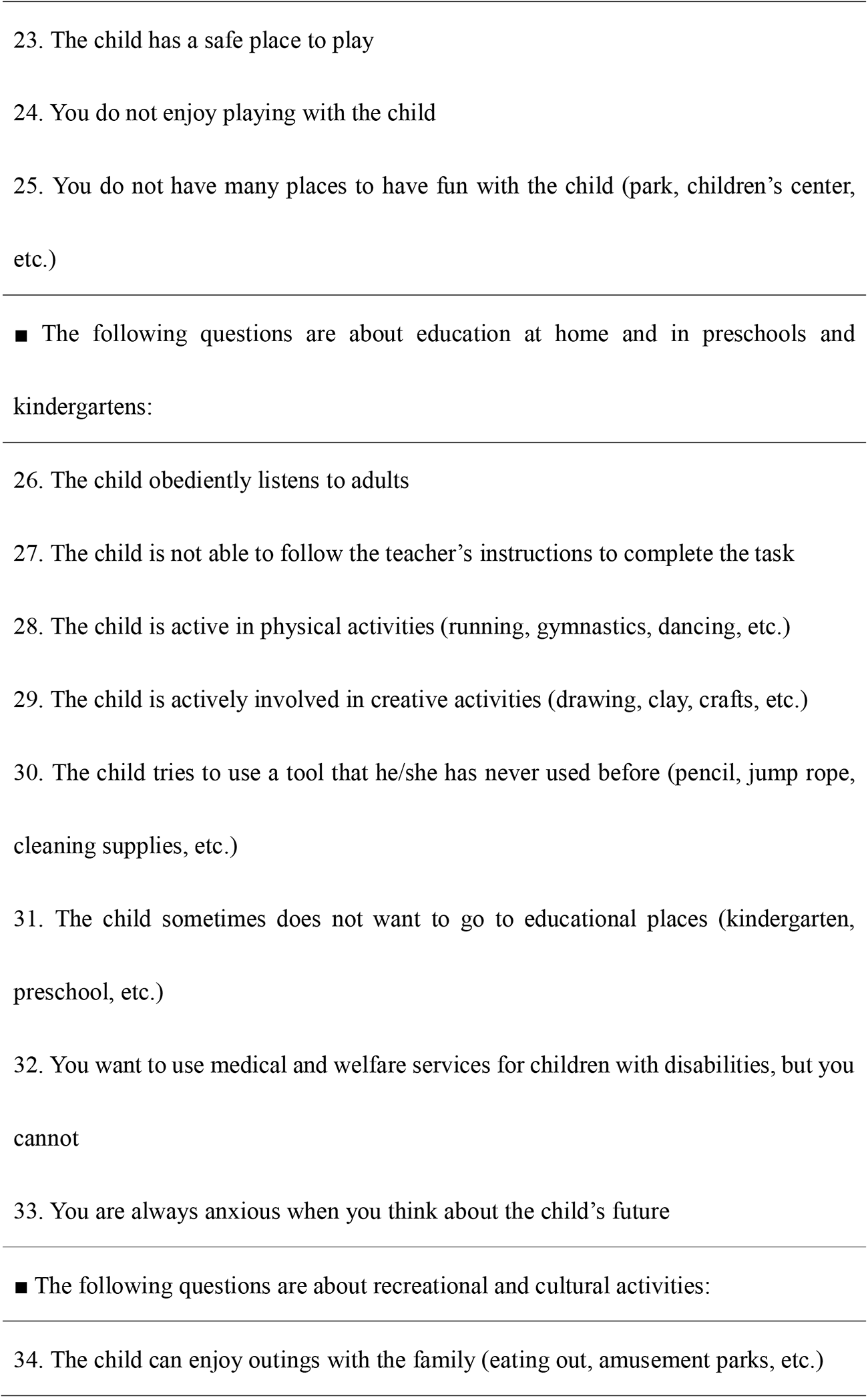
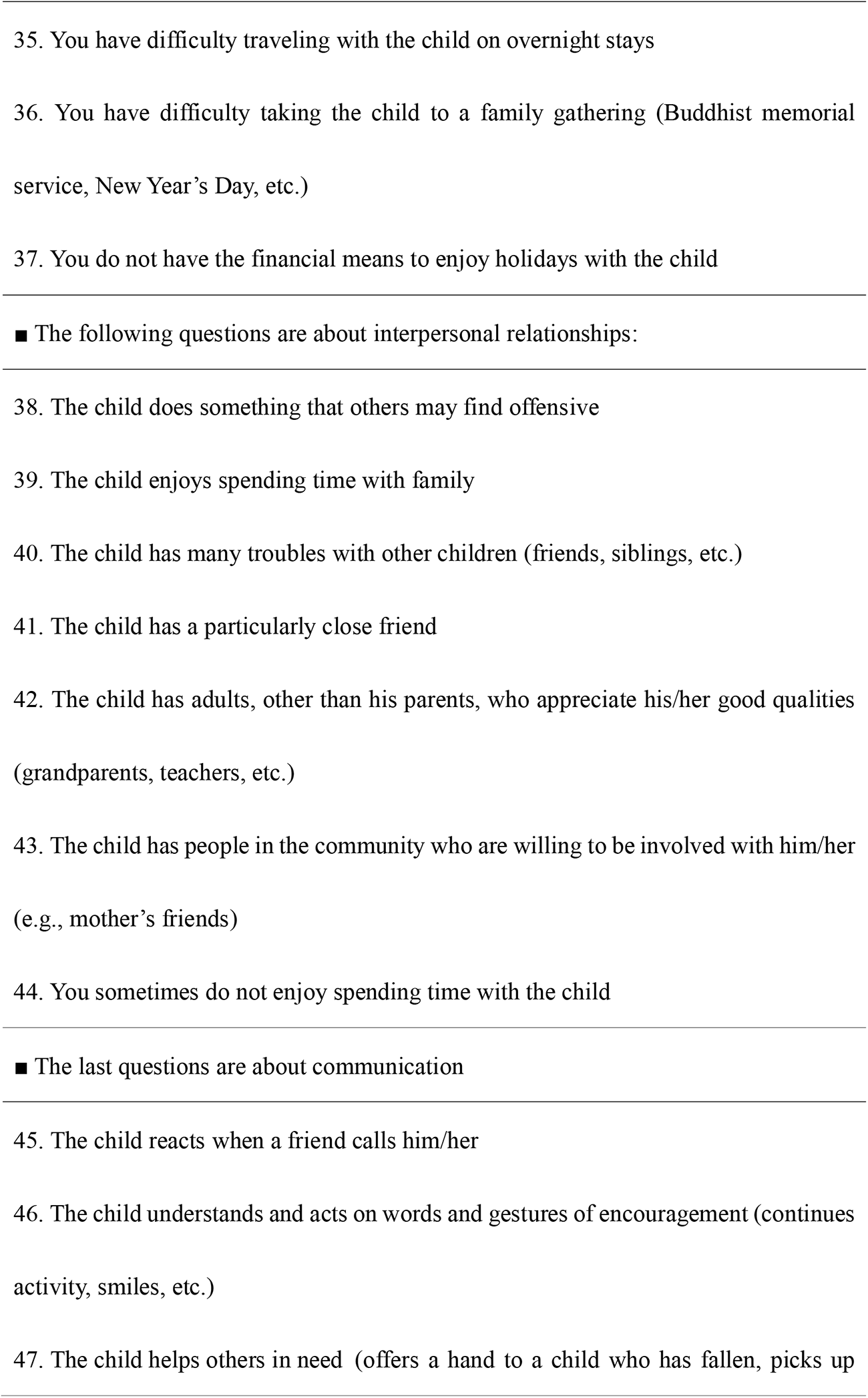
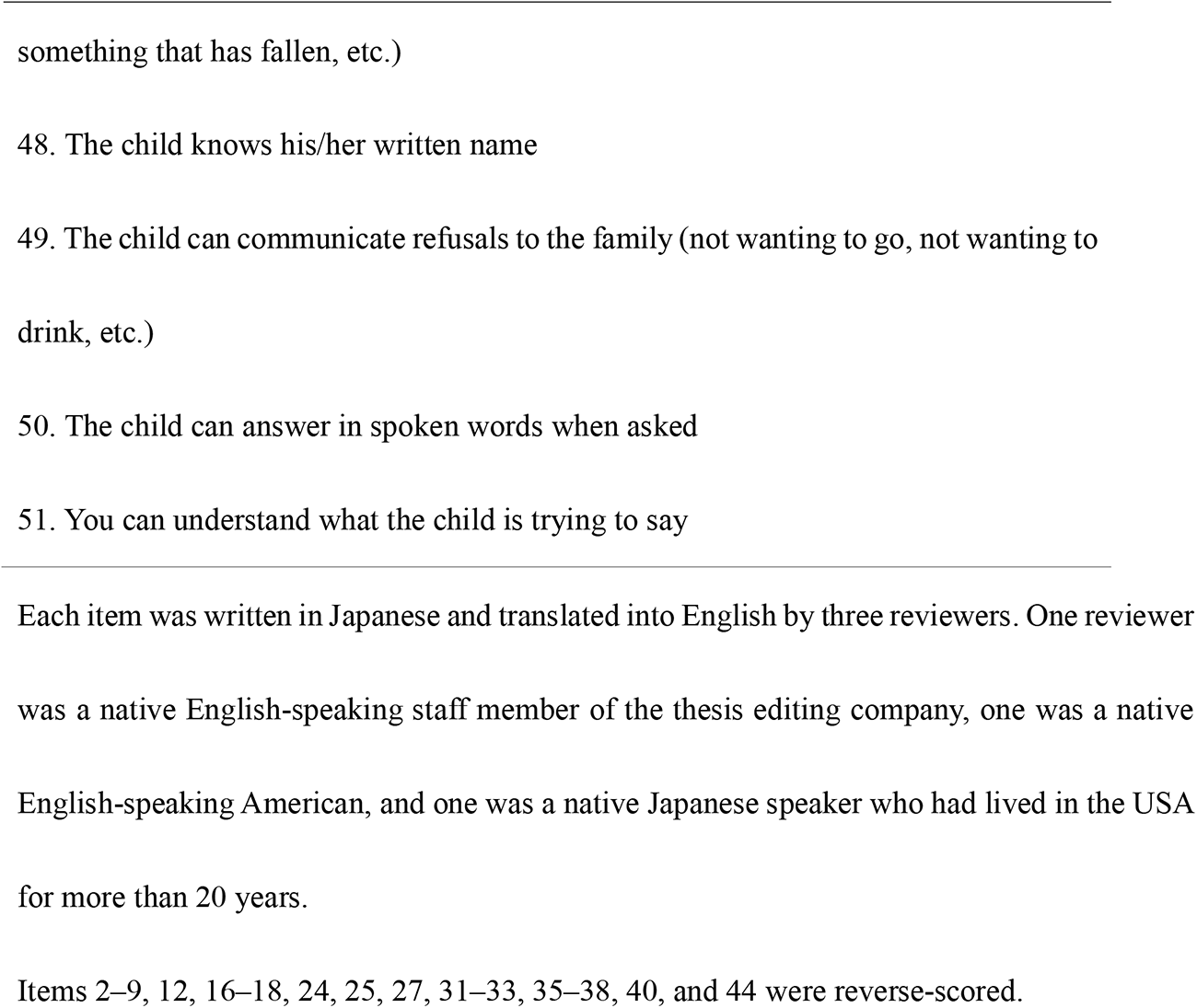
The Participation Questionnaire for Preschoolers: preliminary items and subdomains.

#### Participants

Eight participants were included in the first phase of this study: three “participation” researchers, one occupational therapist, one speech–language–hearing therapist, one clinical psychologist, one certified social worker, and one childcare worker. The inclusion criteria for selecting the participation researchers were that they had published peer-reviewed articles that included the word “participation” in the ICF. Regarding the other professionals, selection was based on them having had at least five years of experience working with children with ASD in their practice. Using a convenience sampling method, participants were approached by e-mail or telephone and informed in advance of the study’s content, purpose, and significance. Of the participants, a total of four were working at universities or other educational/research institutions, one of whom was also working at a clinic, and three were working at child development support facilities or centers, which are typical early support services in Japan. No participant refused to participate in the study. All participants were native Japanese speakers practicing in Japan. Details of participants are presented in Table 2.

**Table 2.**
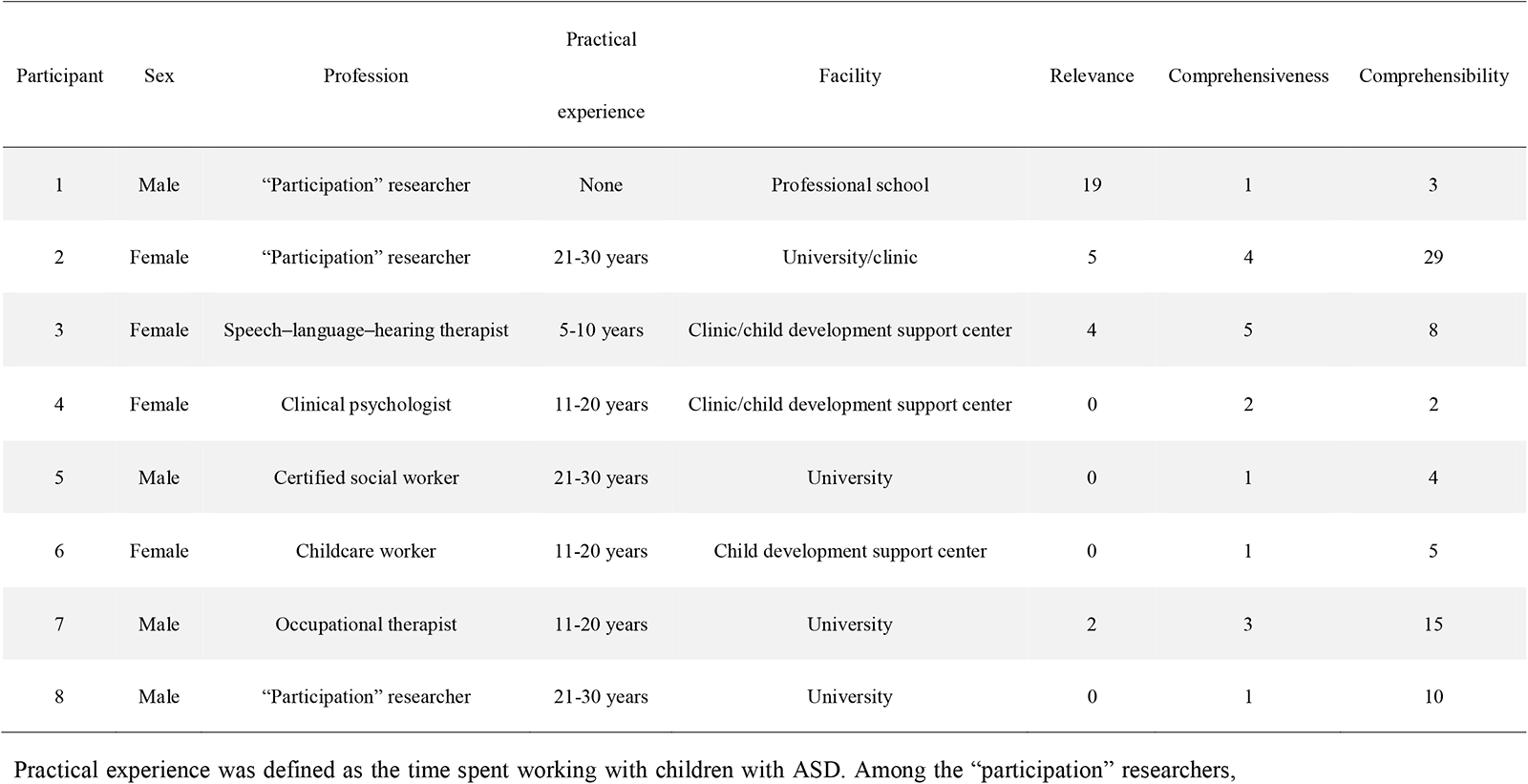
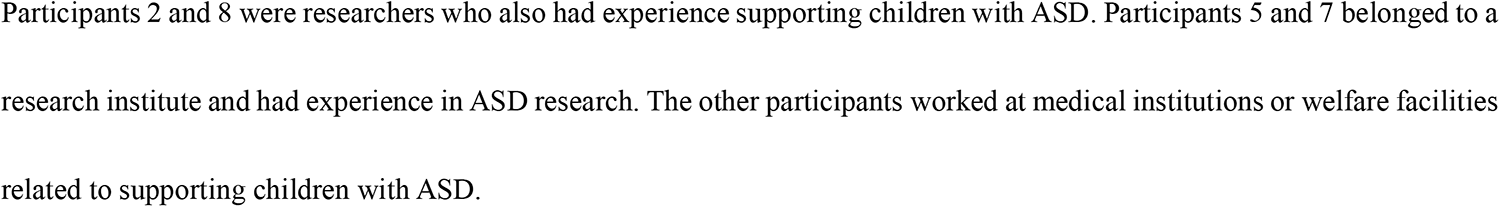
Demographics of the expert participants involved in content validation and the number of codes that appeared.

#### Data collection

Individual semi-structured interviews were conducted with each participant using an online conferencing tool. This format was chosen because there was concern that due to using an online conferencing tool, it would not be possible to have smooth communication among participants in interviews with multiple participants, such as focus group interviews. An interview guide was used for the interviews, asking (1) what were the signs that ASD children’s participation was facilitated or limited, and (2) whether any items were inappropriate or missing from the draft items developed. The first author (TN), a male occupational therapist from a university rehabilitation department with experience in multiple qualitative studies, conducted the interviews in Japanese. The second author (SK), a female occupational therapist with experience in conducting multiple qualitative studies, was also present in the interviews as an interview assistant. The interviews were recorded and transcribed. The average duration of each interview was 93 minutes.

#### Data analysis

A qualitative content analysis was employed [21]. The analysis was conducted first, with coding performed independently by the first and second authors. The unit of analysis was defined as “mentions of item validity.” In this study, validity was defined as any of the following, based on the COSMIN definition of quality guidelines for PROMs [16]:

1. “Relevance” indicates the relevance of the construct being measured (participation), the target population (young children with ASD), the context of use (support for children with ASD), and the appropriateness of the response options and recall period.
2. “Comprehensiveness” indicates the coverage of important concepts for measuring constructs (participation).
3. “Comprehensibility” indicates that respondents have an adequate understanding of the instructions, items, and response options, and that the items are adequately represented.

After each interview, the first and second authors searched the transcripts independently for statements regarding item validity. The statement context was checked to determine whether the statement made a clear reference to the question items. For unclear or ambiguous references, we used the context before and after the statement as a basis for judgment. Next, we determined which item the statement referred to, and assigned an item number to the statement. When a statement referred to more than one question item, multiple codes were extracted from the statement. For example, if a statement referred to both Items 1 and 2, two codes corresponding to each item were extracted. Furthermore, statements that did not refer to a specific item but were related to the questionnaire’s validity, such as those suggesting a lack of items or a need to modify the instructions or questionnaire design, were labeled with the code “other” and extracted. Each code was then deductively labeled for relevance, comprehensiveness, or comprehensibility, based on the aforementioned definition of validity. Finally, these codes were tied to the corresponding item number and participant. For example, a reference to the comprehensibility of the 10^th^ item in the first interview was labeled “1-10-Comprehensibility;” two references to the missing item in the second interview were labeled “2-Other 1-Comprehensiveness” and “2-Other 2-Comprehensiveness.”

After completing the coding, the first and second authors shared their analyses, discussed any discrepancies, and finalized the codes. Following the coding decisions, the two authors read the statements in the generated code; decided whether to modify, delete, or add items (or directives or layouts); and created a draft of revised items. During this process, items from existing scales were sometimes referenced [22, 23]. When new items were modified or added, they were identified as either activity or participation codes with reference to the ICF-CY. This item draft was used in the subsequent round of interviews.

After the interviews were completed, both the first and second authors inductively categorized the codes. These categories grouped the reasons that modifications were deemed necessary, as indicated in the responses. The analysis was conducted separately for codes related to appropriateness and comprehensibility.

### Validation by caregivers

#### Questionnaire design

The draft PQP items as modified in the first phase were used.

#### Participants

This study was conducted with participants who were caregivers of children with ASD. Inclusion criteria were as follows: (1) Participants were caregivers of preschool children aged 36–83 months and with a diagnosis of ASD, (2) the children had no comorbid diagnoses other than neurodevelopmental disorders in the Diagnostic and Statistical Manual of Mental Disorders, Fifth Edition, and (3) the children had an IQ of 50 or higher. During study recruitment, the authors requested referrals from child development support centers and medical institutions in Japan and approached participants by e-mail and telephone. All participants provided oral and written informed consent. No one refused to participate.

Participants were selected to cover variables such as the child’s sex, age in months, comorbid diagnoses, IQ, region of residence, family structure, the caregiver’s sex, and employment status. Specifically, we collected these variables based on reports from caregivers, and when recruiting new participants, we continued to recruit participants in categories that had not yet appeared in one or more variables. The final total number of participants was 11. Details of the participants are presented in Table 3.

**Table 3.**
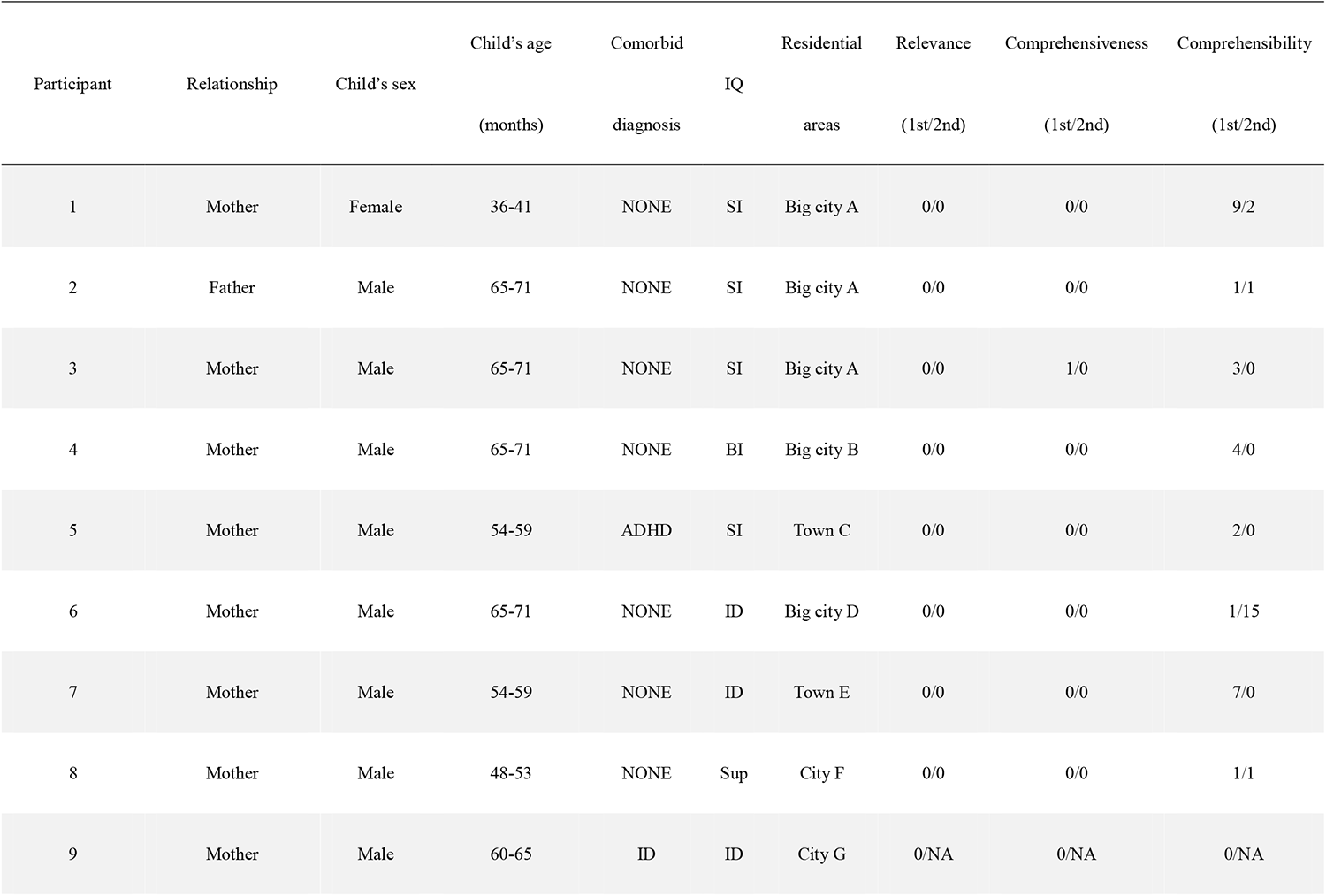
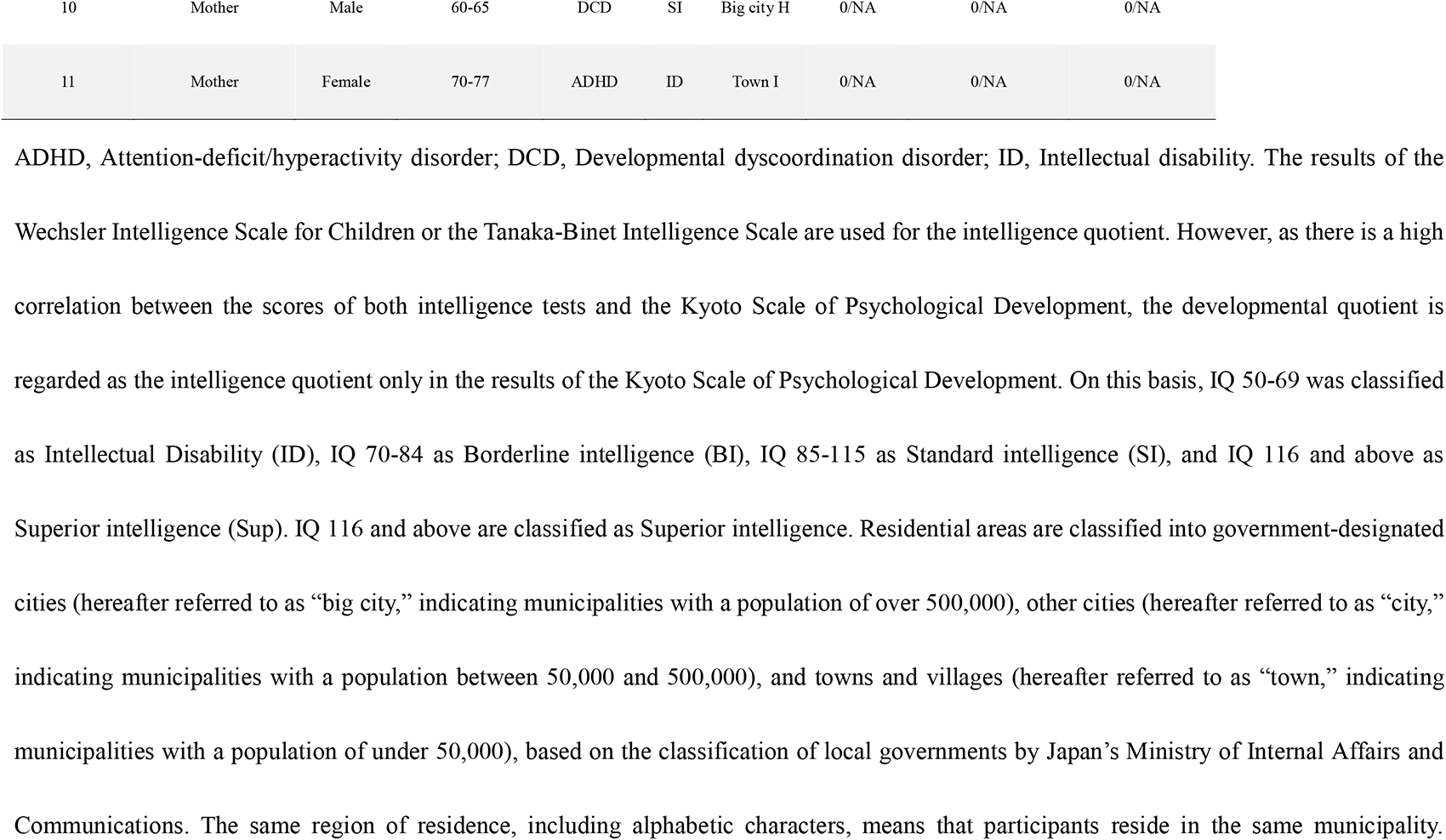
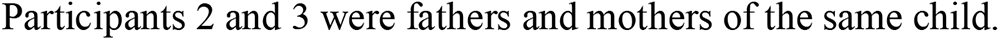
Demographics of caregivers involved in content validation.

#### Data collection

Cognitive and semi-structured interviews were conducted using an online conferencing tool. The interviews were conducted in Japanese by the first author (TN), who received two months of training in cognitive interviewing. In the cognitive interviews, participants were asked to use the “think-aloud method” in combination with the “verbal probing method” [24].

Specifically, the think-aloud method was used for each item in each section of the PQP: “First, please describe your child’s condition;” “Next, please describe your feelings;” and “Finally, please describe your child’s feelings.” If the first author (TN) felt that there was a need for further questions, additional open-ended questions were asked (e.g., “You mentioned that it was difficult to make a judgment, but what specifically was difficult for you?”). In addition, a semi-structured interview was conducted after the cognitive interview. To ensure that the items for understanding participation were covered and that no inappropriate items were included, participants were asked (1) whether the items covered aspects important to the lives of the participants’ children and their families; and (2) whether the drafted items were irrelevant, inappropriate, or offensive to their daily lives. The second author (SK) was also present during the interviews as an interview assistant. The interviews were recorded and transcribed, and either the first or second author took notes for reference in the analysis. In the analysis, participants who needed revisions for any item underwent a second interview to determine if the issues were resolved by the modifications. Data collection was concluded after the interviews with Participant 11, who followed three consecutive participants who did not mention needing revisions, indicating that data saturation had been reached. The average duration of each interview was 33 minutes.

#### Data analysis

The same analysis was performed as in Phase 1.

## Results

### Content validation by experts

A total of 120 changes were made to the proposed PQP questions, reducing the final number of questions from 51 to 35. The changes comprised 80 modified questions, 28 deleted questions, and 12 added questions. The sub-domain most frequently reported was “education,” (35 times; 29.2%), followed by “self-care” (23 times; 19.2%), “interpersonal interaction and relationships” (15 times; 12.5%), “community, social and civic life” (14 times; 11.7%), “engagement in play” (13 times; 10.8%), “domestic life” (8 times; 6.7%), “communication” (7 times; 5.8%), and “mobility” (4 times; 3.3%). One modification (0.8%) was also made to the overall design of the questionnaire. Table 2 shows the number of occurrences of the relevance, comprehensiveness, and comprehensibility codes in each interview.

#### Relevance

There were 30 relevance codes, accounting for 25% of all codes. When the relevance codes were inductively categorized according to their reasons, the most frequently mentioned code was “clarifying the difference between activity and participation” (n = 17, 56.7%). Approximately 82.4% of these codes were attributed to Participant 1 (a “participation” researcher). It was ascertained that all questions within the communication and mobility domains pertained more to activity than participation, leading to the deletion of these two domains. Additionally, Participant 1 referenced “clarifying the difference between environment and participation” (n = 5) 16.7% of times.

> I think the sub-domain of communication often asks about skills, so, well, if you ask if it’s about participation, I will say it’s not really. (Participant 1)

In addition, “the meaning of the question changes depending on the context” (n = 7) was mentioned by Participants 2 (“participation” researcher) and 3 (speech–language–hearing therapist), accounting for 23.3% of the total. Another code, which indicated items that highlighted caregivers’ concerns, was identified: “raises concerns among caregivers.” Codes for appropriateness appeared in 93.3% of the total responses from Participant 3, after which only two codes appeared from Participant 7.

#### Comprehensiveness

There were 14 codes for comprehensiveness, accounting for 11.7% of all codes. Half of the codes (n = 7) were situated within the education domain.

> I think it’s important for children of the same age to have opportunities; for example, to get together in a group like during a music festival or a play or a Christmas party and enjoy the atmosphere. Whether it’s possible to share time and place … I feel like this is something important for participation. (Participant 5)

In response, a new item was added in the education domain: “Has opportunities to enjoy reading with children of the same age” (later revised to “Enjoys class activities in cooperation with children of the same age”).

#### Comprehensibility

There were 76 comprehensibility codes, accounting for 63.3% of all codes. Comprehensibility codes were inductively classified as “question structure is difficult to understand” (n = 41) for 46.1%, “topic is unclear” (n = 39) for 43.8%, “answer is difficult to determine” (n = 8) for 9.0%, and “instructions are difficult to understand” (n = 1) for 1.1% (13 codes were overlapped into two classifications). In particular, in the interview with Participant 2, it was suggested that the items, which had been categorized into eight domains according to the ICF activity and participation codes, be presented in the following three question formats: “answer the following questions about your child,” “describe how you feel,” and “answer the question about your child’s feelings.”

> The subjects have been repeatedly “this child is” and “you are.” This item comprises two types of questions: “A question to answer about the child as understood from the mother’s perspective” and “A question for the mother to speculate from the child’s perspective.” We must consider that changing the subject requires effort for people with tendencies of neurodevelopmental disorders, and it could be burdensome for them to answer. (Participant 2)

With this change, the items that had been arranged as eight domains were rearranged into the three question formats described above.

#### Content validation by caregivers

A total of 49 changes were made to the PQP items, resulting in the final number of questions changing from 35 to 36. There were 48 modifications, zero deletions, and one addition. Interpersonal interaction and relationships were modified 20 times (40.8%); education, 13 times (26.5%); engagement in play, eight times (16.3%); community, social, and civic life, four times (8.2%); self-care, two times (4.1%); and modifications regarding response options, two times (4.1%). No modifications were made to “domestic life.” The number of occurrences of the relevance, comprehensiveness, and comprehensibility codes in each interview is shown in Table 3; and the revised items, after the interviews were completed, are shown in Table 4.

**Table 4.**
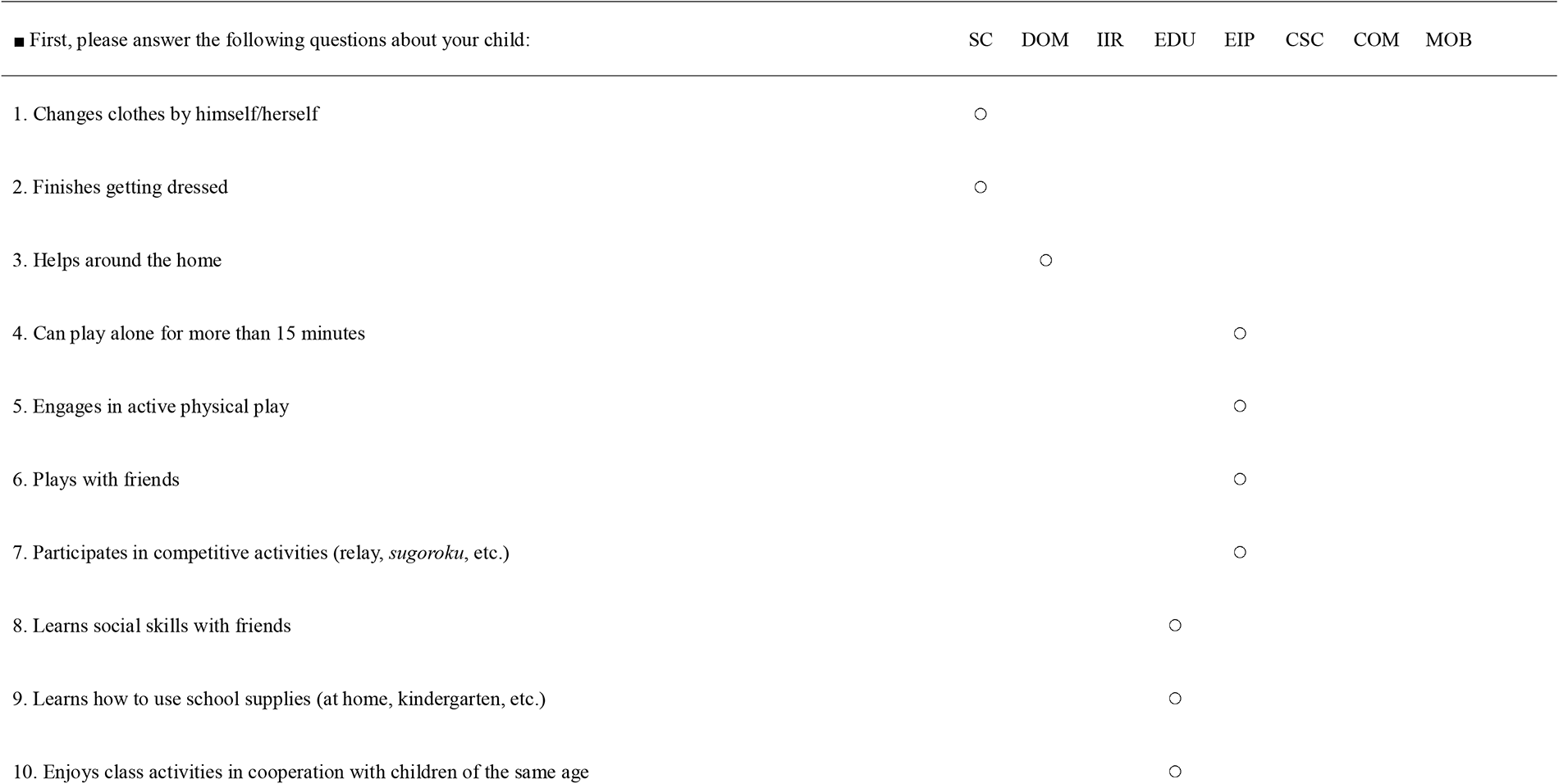
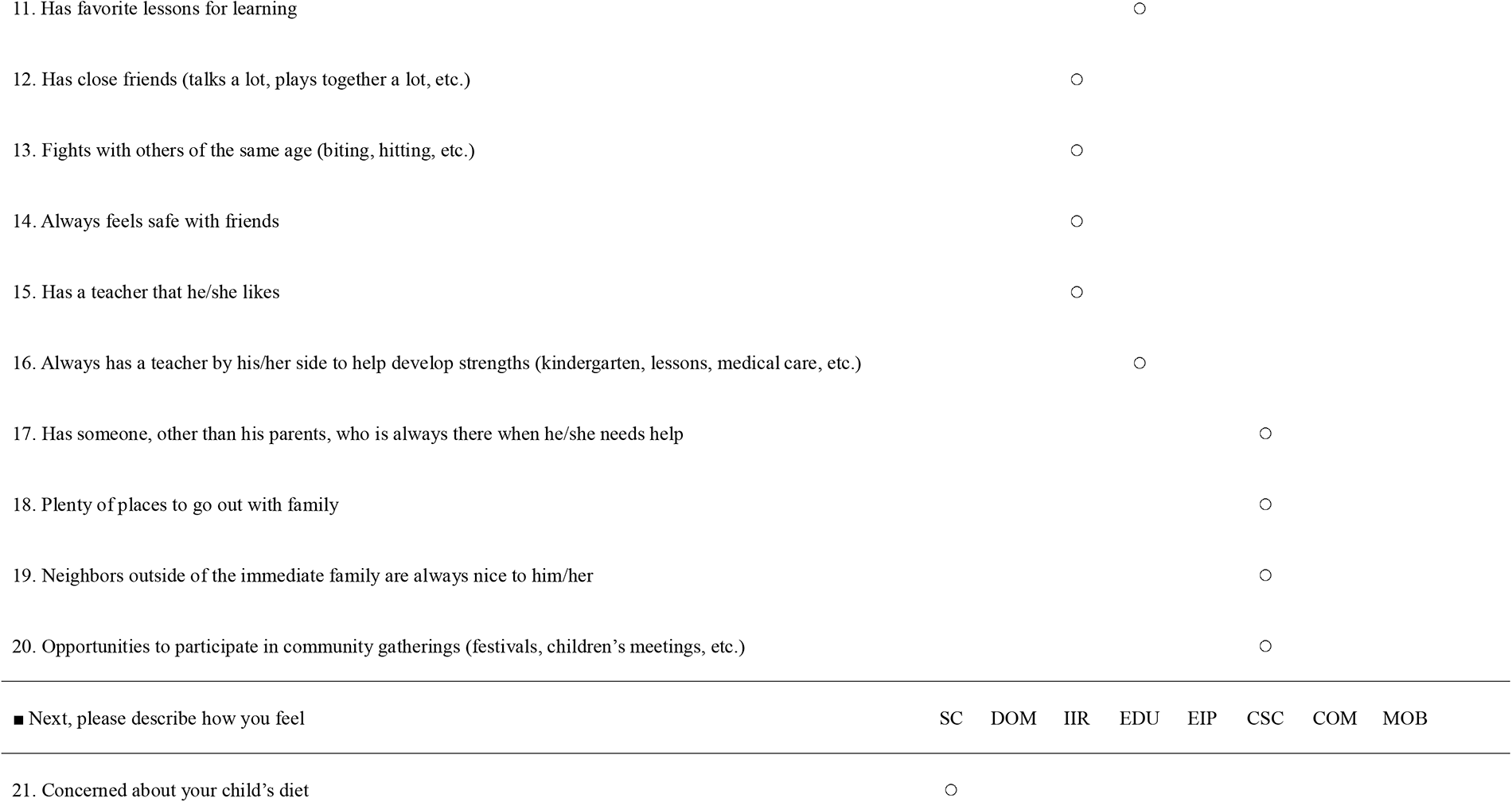
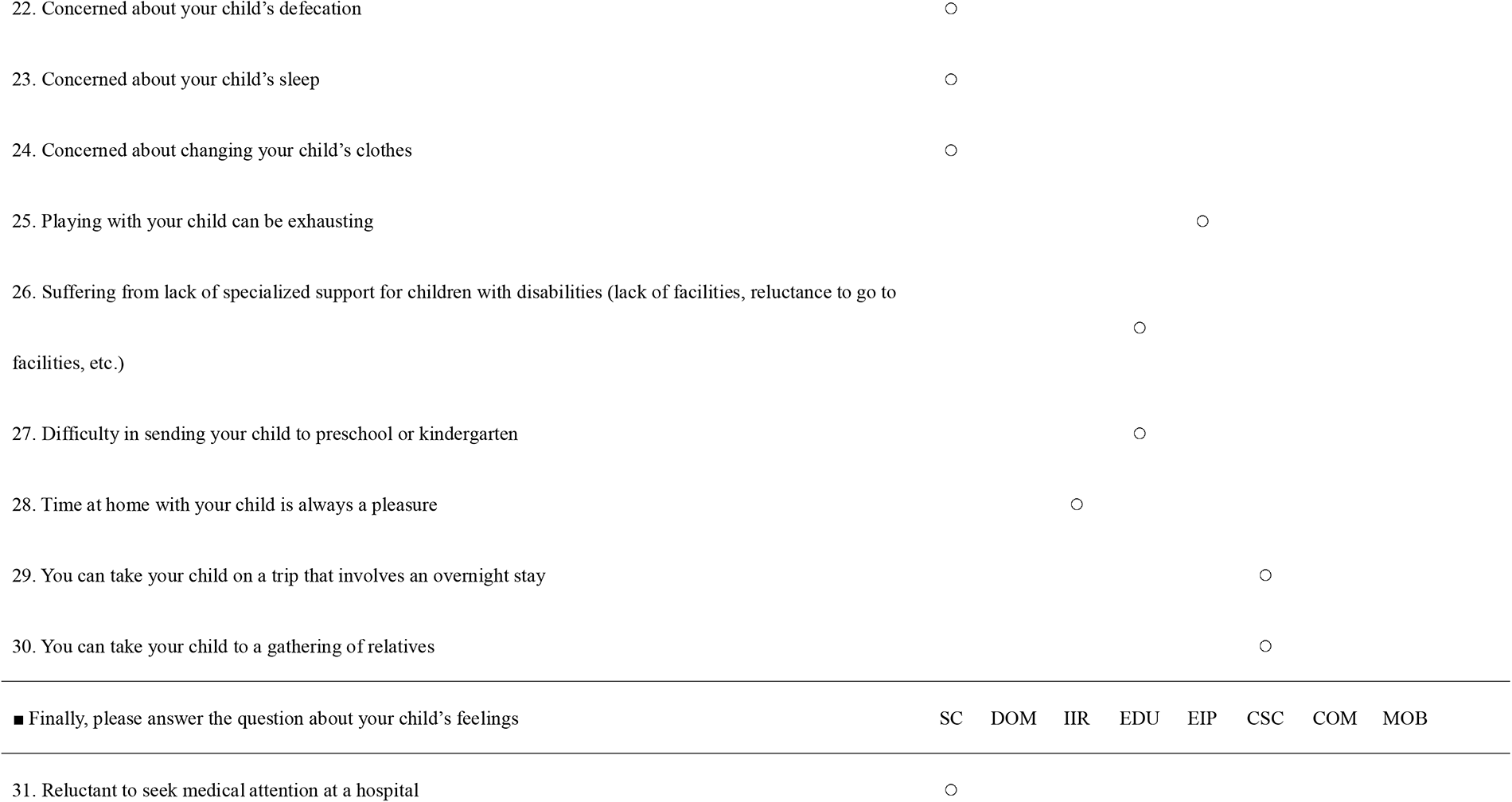
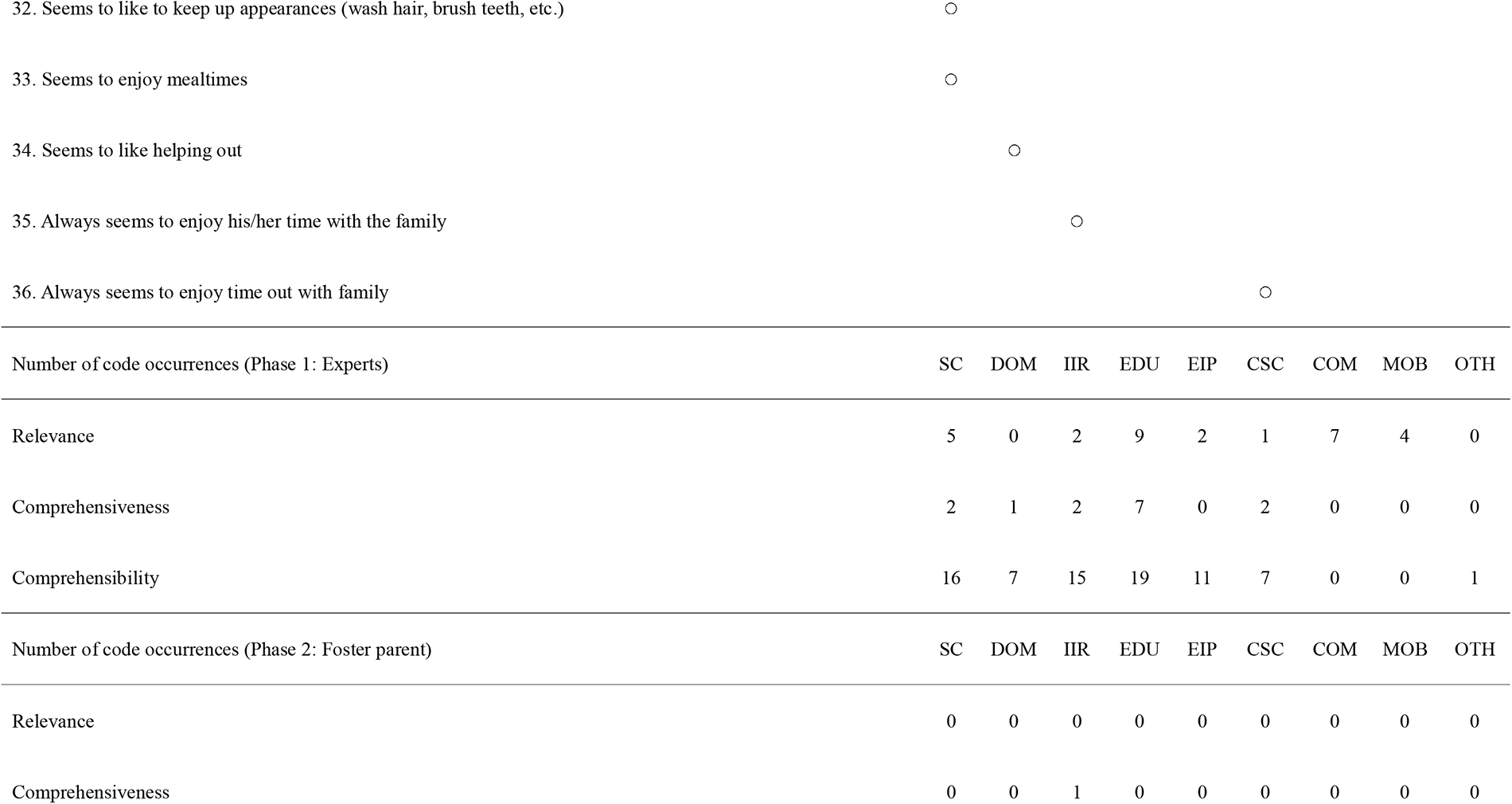
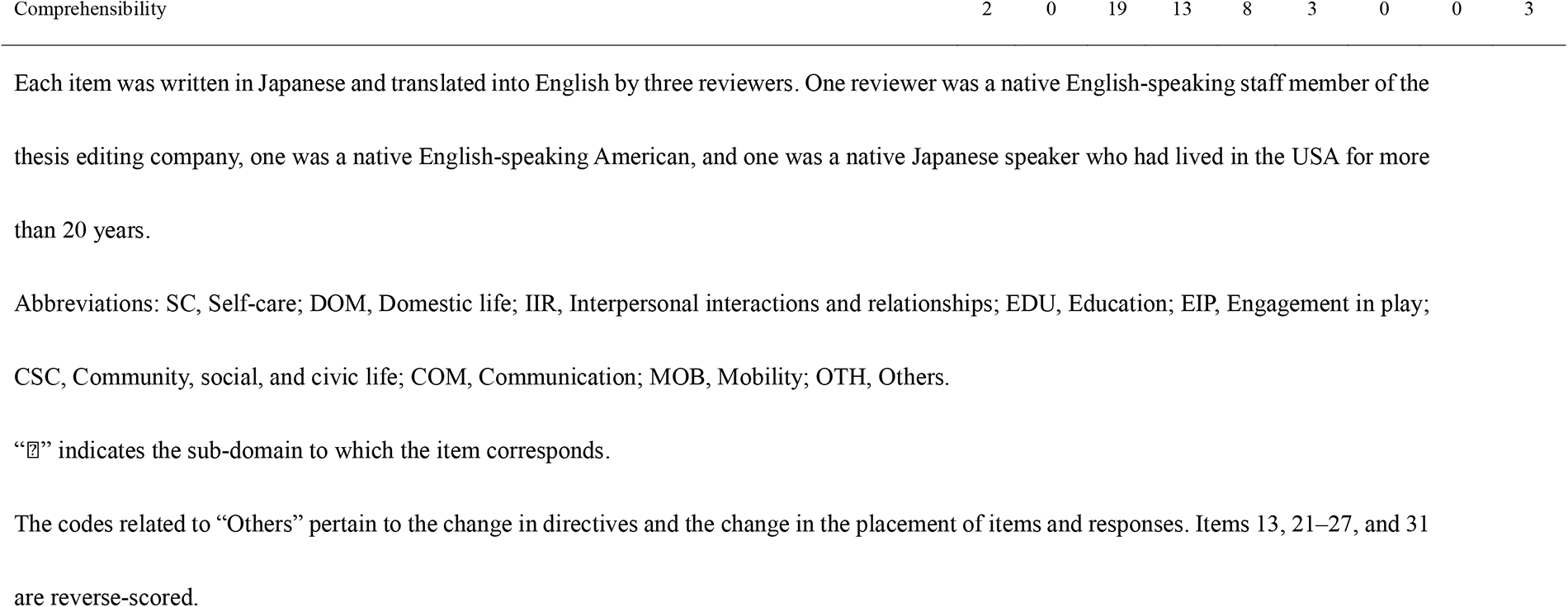
Items after caregivers’ content validation and the number of codes for each sub-domain.

#### Relevance

No issues of relevance were reported in the interviews. All participants stated that the items were relevant to the child, and the questions were not deemed sensitive for respondents to answer.

#### Comprehensiveness

Only one code for comprehensiveness was reported, accounting for 2.1% of the total. A mother of a child within the highest quartile for age in the study noted that there were few questions about relationships with friends, commenting:

> “I have no problem changing clothes and daily habits, and I am worried about what is going on with their friends, so I thought it would be okay if I had some more questions like that.” (Participant 3)

A new item—“Has close friends (talks a lot, plays together a lot, etc.)”—was added in response.

#### Comprehensibility

Regarding comprehensibility, there were 48 references to question ambiguity and misleading item order, accounting for 98.0% of the total. The codes were inductively categorized as follows: “topic is unclear” (41.7%, n = 20), “question is difficult to recall” (18.8%, n = 9), “answer is difficult to determine” (16.7%, n = 8), “question structure is difficult to understand” (8.3%, n = 4), and “others” (14.9%, n = 7). The “others” included the content of the instructions, the order in which the items were presented, and the placement of the response options (applicable options on the left and non-applicable options on the right). For example, although Participant 6 talked about her child’s challenging behavior at mealtime in the semi-structured interview, she answered “agree” to the item “family time seems to be fun.” However, when she was probed by the interviewer about whether she rarely had fun during family time, she replied as follows.

> When he doesn’t get his way, there are quite a few times when he gets angry … But the joyful look on my child’s face is quite impressionable to me, so I ended up choosing “agree” for this question. (Participant 6)

This item was judged to be a “question difficult to recall” and was revised to “always appears to enjoy his time with his family.”

## Discussion

### PQP item development and characteristics

This study aimed to extend the age range targeted by the PQP and to develop new items suitable for a broader spectrum of children. The newly developed PQP items offer three primary advantages. First, they enable the comprehensive collection of participation data through a questionnaire, which is essential. The PQP was validated for its relevance, comprehensiveness, and comprehensibility based on interviews with eight experts and eleven caregivers of children with ASD until data saturation was achieved. Looking forward, if the development of this scale is completed and it allows for assessing comprehensive participation without requiring trained professionals, it could greatly improve the feasibility of conducting large-scale surveys. Additionally, this could establish a foundation for gathering evidence about the participation of children with ASD.

Second, the newly developed items reflect the perspectives of both professionals and caregivers, highlighting the key challenges faced by children with ASD, such as difficulties performing activities of daily living and accessing specialized services [2]. Disability-specific measurement tools may be better suited to capture the information required for intervention and monitor intervention effectiveness than generic instruments [8, 25]. The PQP could, therefore, be a useful tool for occupational therapists to assess and monitor the participation of children with ASD during interventions.

Third, this tool facilitates the assessment of a wider age range of children with ASD than the previous PQP items enabled. The need for early intervention for children with ASD has been widely recognized, and intervention implemented from infancy is becoming more common in high-income countries [2]. Therefore, the PQP’s expanded age range increases its accessibility for practical use. It could also contribute to the feasibility of research on the early childhood participation of children with ASD, such as by enabling longer-term longitudinal studies.

### Limitations and future research directions

The development of measurement tools requires item development, scale development, and the verification of reliability and validity [26]. To make the PQP usable in practice and research, these steps must be implemented in the future. Particularly, the selection of questionnaire items will require item reduction analysis for scale development, and the validation of content validity needs to be conducted separately after scale development. COSMIN recommends validating content validity using qualitative research and also recommends the use of quantitative approaches such as the Delphi method alongside the qualitative methods as beneficial [16].

This study has some limitations. Between 2020 and 2021, when the data were collected, many municipalities in Japan implemented travel restrictions owing to the COVID-19 pandemic [27]. This could have limited many people’s diversity of events and travel-related items. To address this issue, it may be useful to reexamine the content validity of the data by comparing it with data collected during the non-pandemic period. In addition, the data collected from caregivers included only participants who used child development support services and medical institutions. Therefore, the participation status of children who did not use these services may not have been adequately assessed. In future studies, it will be important to assess the applicability of the PQP to children who do not use these services. This would enable confirmation that the PQP adequately reflects the participation status of all children with ASD. Finally, the content of the newly developed PQP items was validated only in the unique cultural and institutional environment of Japan. Cross-cultural validation is required to use the PQP in countries other than Japan.

## Conclusion

This study expands the target age range of the PQP—a disability-specific participation measurement tool for children with ASD—to 38–83 months and develops new items. The updated version of the PQP has two unique features: (1) it provides a comprehensive assessment of participation in a questionnaire format, and (2) it includes items specific to the challenges faced by children with ASD.

## Acknowledgments

The authors express their gratitude to medRxiv for hosting our initial findings, available at https://www.medrxiv.org/content/10.1101/2023.08.22.23294206v1. We also thank Erik and Satomi Phillips for their assistance with the English translation of the items.

## Data Availability Statement

The data that support the findings of this study are available upon reasonable request but are not publicly accessible due to privacy or confidentiality concerns.

